# Performance characteristics of the ID NOW COVID-19 assay: A regional health care system experience

**DOI:** 10.1101/2020.06.03.20116327

**Authors:** Mohiedean Ghofrani, Mary T. Casas, Robert K. Pelz, Catherine Kroll, Natalie Blum, Scott D. Foster

## Abstract

**Objectives:** We compared the Abbott ID NOW COVID-19 point-of-care test (POCT) with polymerase chain reaction (PCR)-based methods to assess the claimed sensitivity and specificity of POCT and to optimize test utilization in our regional health care system.

**Methods:** Assuming PCR to be the gold standard, we used a convenience sampling of mostly symptomatic COVID-19 suspect hospital patients who had already been tested for internal validation and guideline development purposes by both PCR and POCT to calculate the sensitivity and specificity of POCT with Clopper-Pearson 95% confidence intervals (CI).

**Results:** During the study period, 113 paired patient samples met eligibility criteria. The sensitivity of POCT in this population was calculated to be 94.1% [CI 71.31-99.85%] and the specificity was 99.0% [CI 94.33-99.97%].

**Conclusions:** Based on the lower sensitivity of POCT and the estimated prevalence of COVID-19 in our symptomatic and asymptomatic hospital patients, we recommend a two-pronged testing approach in which COVID-19 suspect patients are tested by the more sensitive PCR, while asymptomatic patients with a low pre-test probability of infection are tested with POCT supplemented by PCR confirmation of positive results. Furthermore, isolation decisions should not be based on POCT results alone.

## Introduction

In December 2019 a cluster of pneumonia cases was reported in Wuhan, China, that was later found to be caused by a novel coronavirus eventually named severe acute respiratory syndrome coronavirus 2 (SARS-CoV-2)^1^. Within the span of a few weeks, infections with the virus rapidly spread throughout the world. By March 2020, the World Health Organization designated coronavirus disease 2019 (COVID-19) a pandemic, and the United States declared it a National Emergency^2^.

Similar to influenza, person-to-person spread of COVID-19 is thought to mainly occur through respiratory droplets; at the time of this writing, airborne transmission under normal conditions is a subject of debate^3,4^. This necessitates, at a minimum, the use of contact and droplet precautions including appropriate isolation and personal protective equipment (PPE) when caring for suspect patients, as well as airborne precautions during aerosol-generating procedures. However, patient isolation is a resource-intensive task, and PPE is in limited supply in most health care settings and communities at large. Efficient resource management would therefore dictate that isolation and PPE only be used for patients who are truly infected in order to avoid using them in those who do not pose a risk of spreading the virus. Given the varied presentations of COVID-19 in terms of incubation period^5^, symptomatology^6,7^, and disease severity^8^ (including asymptomatic infections^9^), and the reported infectivity of even asymptomatic patients^10^, accurate laboratory testing to reliably identify infected individuals has become an integral part of managing COVID-19 suspect patients and fighting the spread of SARS-CoV-2 in the community.

To meet this critical need for laboratory testing, beginning on February 4, 2020, the U.S. Food and Drug Administration (FDA) relaxed its normal process for approval of in vitro diagnostics by issuing Emergency Use Authorizations (EUAs) so clinical laboratories could quickly introduce new tests to detect SARS-CoV-2 in patients suspected of COVID-19^11^. EUA status was first granted to a number of polymerase chain reaction (PCR)-based tests that were authorized to be used in laboratories certified to perform moderate- and high-complexity testing under the Clinical Laboratory Improvement Amendments of 1988 (CLIA)^12^. PCR-based tests have since become the “gold standard” for laboratory detection of SARS-CoV-2^13,14^.

On March 27, 2020, Abbott Diagnostics Scarborough, Inc. (Scarborough, ME) received EUA for the ID NOW COVID-19 assay as a point-of-care test (POCT) to be performed not only in clinical laboratories certified to perform moderate- and high-complexity testing but also in patient care settings outside of the clinical laboratory that operate under a less stringent CLIA Certificate of Waiver or Certificate of Compliance^15^. The ID NOW COVID-19 assay differs from conventional PCR in that it utilizes an isothermal nucleic acid amplification technology for qualitative detection of the RNA-dependent RNA polymerase (RdRp) gene of SARS-CoV-2^16^. It typically produces positive results in 5 minutes and negative results in 13 minutes, compared to the turnaround time of PCR, which for our health care system is measured in hours to days depending on ordering location and performing laboratory.

Beginning on April 10, 2020, our 10-hospital regional health care system decided to include the ID NOW COVID-19 POCT as an in-house laboratory testing option to complement our established process of PCR testing at commercial, State Public Health, and academic laboratories. In the course of internally validating POCT and developing guidelines to assist ordering providers in choosing either PCR or POCT testing, paired samples were collected from hospital patients to evaluate POCT performance characteristics as claimed by the manufacturer.

## Materials and Methods

This study was approved by our system institutional review board and conducted within our health care system composed of 10 hospitals of various sizes and numerous clinics serving suburban and rural communities in three states. No funding was received for this study. Paired samples from patients who had already been tested for purposes of internal validation and guideline development by both send-out PCR and in-house ID NOW COVID 19 POCT from April 6 through April 21, 2020, were considered for inclusion in the study.

Paired PCR and POCT samples had already been collected per manufacturer instructions by nasopharyngeal swab or mid-turbinate nasal swab. The majority of these patients had symptoms suspect for COVID-19, including patients who had a PCR sample collected close to the time of presentation followed by a re-swab for POCT, and those who were already known to be PCR-positive and the residual nasopharyngeal specimen was tested by POCT. A handful of patients did not have symptoms of COVID-19, but because they tested positive by POCT on admission they were re-swabbed for PCR confirmation per institutional guidelines. Since this was a convenience sampling of mostly symptomatic patients with a bias to re-swab patients with a known positive test result, this enriched cohort cannot be used to estimate overall COVID-19 prevalence in our hospital inpatient population.

Nasopharyngeal swabs collected for PCR were placed in universal viral transport (UVT) medium, while nasal and nasopharyngeal swabs collected for POCT were either submitted directly for testing (dry swab) or in UVT. PCR samples were sent, depending on local logistical considerations, to either one of two commercial laboratories, one of two State Public Health laboratories, an academic medical center, or tested in-house. Although the ID NOW COVID-19 assay is authorized for testing in patient care settings outside of the clinical laboratory, all POCT testing was performed in our hospital laboratories by laboratory personnel.

De-identified data were recorded in an Excel spreadsheet (Microsoft, Redmond, WA). Recorded data fields for each PCR sample included collection date, send-out laboratory where test was performed, and PCR result. Recorded data fields for each POCT sample included collection date, specimen source, transport medium (if any), hospital laboratory where test was performed, and POCT result. If the paired PCR and POCT specimens were collected more than 3 days apart, they were not considered concurrent and excluded from the study.

Assuming PCR to be the gold standard, we calculated the sensitivity and specificity of POCT in our study population and determined Clopper-Pearson 95% confidence intervals for the two.

## Results

Based on the above-mentioned inclusion and exclusion criteria, paired samples from 113 unique patients were eligible for this study. The distribution of collection time differences of the paired samples is illustrated in Figure 1. Of the 113 PCR tests, 60 (53.1%) were performed at one of two commercial laboratories (Quest Diagnostics or a local laboratory), 46 (40.7%) were performed at one of two State Public Health laboratories, 5 (4.4%) were performed at an academic medical center, and 2 (1.8%) were performed in-house. Seventeen (15.0%) of these samples were PCR-positive and 96 (85.0%) were negative.

**Figure 1.**
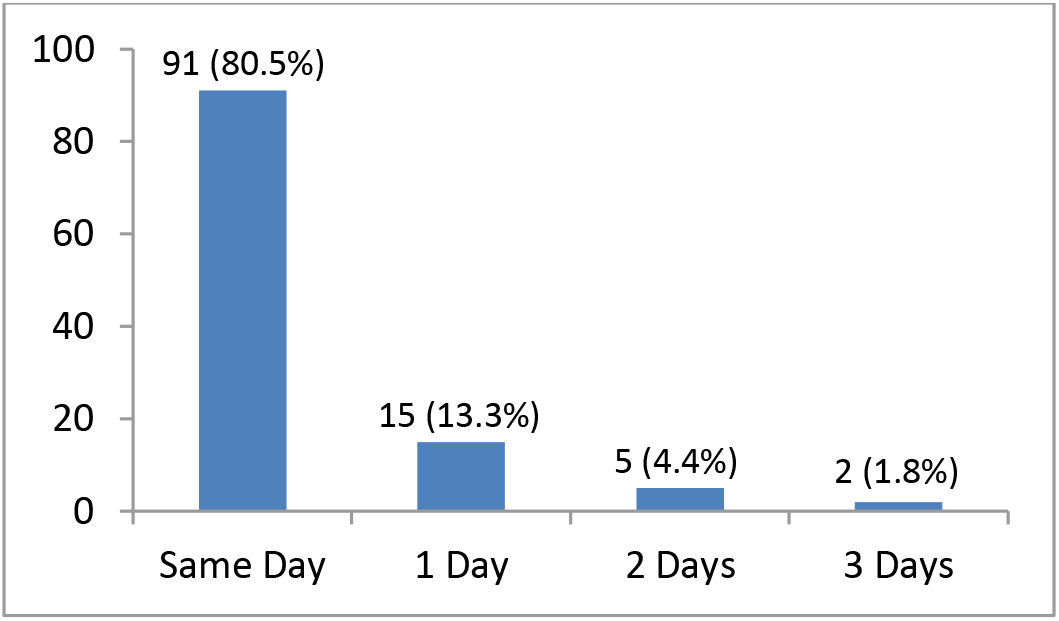
Distribution of collection time differences of 113 paired samples.

All 113 POCTs were performed in one of nine hospital laboratories within our health care system. Of these, 58 (51.3%) were nasal swabs, 33 (29.2%) were nasopharyngeal swabs, and in 22 cases (19.5%) the sample source had not been recorded. POCT swabs were tested directly (dry swabs) in 58 cases (51.3%), transported in UVT before testing in 26 cases (23.0%), and in 29 cases (25.7%) the type of transport medium, if any, had not been recorded.

Assuming PCR to be the gold standard, 16 of the 17 PCR-positives were also positive by POCT, while one was false negative. This one false negative POCT sample was a thawed residual nasopharyngeal specimen in UVT that was known to have tested positive by PCR. Of the 96 cases negative by PCR, 95 were also negative by POCT, while one was false positive. The false positive POCT sample was a nasopharyngeal specimen in UVT. The calculated sensitivity of POCT in this series of paired samples was 94.1%, with a Clopper-Pearson 95% confidence interval (CI) of 71.31% to 99.85%. The specificity of POCT was 99.0%, with a 95% CI of 94.33% to 99.97%. These results are summarized in Table 1.

**Table 1.** Results of concurrent PCR and POCT.

|  | PCR-positive | PCR-negative | Total |
| --- | --- | --- | --- |
| POCT-positive | 16 | 1 | 17 |
| POCT-negative | 1 | 95 | 96 |
| Total | 17 | 96 | 113 |

Calculated sensitivity = 94.1% [CI 71.31-99.85%]
Calculated specificity = 99.0% [CI 94.33-99.97%]

## Discussion

Although the sensitivity and specificity of assays for the detection of SARS-CoV-2 have not been systematically evaluated, PCR testing is currently the gold standard to which the performance of other testing methodologies is commonly compared. The ID NOW COVID-19 assay is claimed by the manufacturer to have an analytical sensitivity of 95% at a limit of detection of 125 genome equivalents/mL based on a study of 20 contrived positive samples (as described in the product insert) and a specificity of at least 97% (verbal communication with the manufacturer).

At the time of this writing, we have identified only a few peer-reviewed clinical studies comparing the performance of the Abbott ID NOW COVID-19 assay with PCR. In one study of 96 remnant clinical specimens, 85 provider-collected nasopharyngeal swabs in universal transport medium (UTM) and 11 self-collected nasal swabs in saline were tested by the Abbott ID NOW COVID-19 assay and compared to a modified Centers for Disease Control and Prevention (CDC) PCR method; positive percent agreement (PPA) was reported to be 94% [CI 87-98%]^17^. In another study of 108 nasopharyngeal swabs collected from symptomatic patients and transported in UTM, 58 samples tested positive and 50 tested negative by the reference standard (Panther Fusion SARS-CoV-2 PCR assay); PPA of the ID NOW COVID-19 assay was 87.7% [CI 76-95%] and negative percent agreement (NPA) was 100% [CI 93-100%]^18^.

Of note, the original ID NOW COVID-19 product insert allowed for elution of swabs in saline or viral transport medium in addition to dry swabs. However, on April 21, 2020, the instructions were revised to direct swabbing only, as it was found that dilution in transport medium may result in decreased detection of low positive samples^19^. In one study, 524 dry nasal swabs were tested directly or transported in sterile tube before testing by the ID NOW COVID-19 assay, consistent with the updated manufacturer’s instructions, and compared to paired nasopharyngeal swabs transported in viral transport media tested with the Abbott m2000 PCR method; the PPA of ID NOW to PCR was 74.73% [CI 67.74-80.67%] and the NPA was 99.41% [CI 97.64-99.89%]^20^.

Given the EUA status of COVID-19 in vitro diagnostics and the limited availability of validation data when we introduced the ID NOW COVID-19 assay into our health care system’s laboratory testing menu, we were asked by our providers to perform a paired sample study of at least 100 patients to internally validate the claimed sensitivity and specificity of POCT. Our study showed the 94.1% sensitivity of POCT in our patient population to be slightly less than the 95% sensitivity stated by the manufacturer. However, the stated 95% sensitivity falls within the Clopper-Pearson 95% confidence interval calculated in our patients. POCT showed a very high specificity of 99%, comparable to PCR and in agreement with the manufacturer’s claimed specificity of over 97%.

Viral load is known to change during the clinical course of COVID-19. In a study of temporal patterns of viral shedding in 94 patients who were, at most, moderately ill, 414 throat swabs were collected from symptom onset up to 32 days later for quantitative PCR testing^21^. High SARS-CoV-2 viral loads were detected soon after symptom onset, followed by gradual decline to the detection limit by about day 21, with no obvious difference in viral loads across sex, age and disease severity. The authors inferred from this observation that infectiousness peaked on or before symptom onset. In another study of symptomatic COVID-19 patients that included daily measurements of SARS-CoV-2 by PCR, viral loads were found to already be on the decline at the time of presentation in the majority of cases^22^. This is in contrast to SARS-CoV, where peak RNA concentrations were usually detected 7 to 10 days after presentation^23,24^. Given the above evidence we therefore chose to exclude paired samples that were collected more than 3 days apart in order to avoid negative result bias due to naturally declining viral loads.

## Conclusions

Due to limited access to testing in early stages of the pandemic, our health care system decided to utilize both send-out PCR and in-house POCT to ensure testing of as many of our hospital admitted patients as possible. In order to optimize the accuracy and timeliness of test results that would ultimately guide decisions regarding treatment, isolation and PPE use, we decided to use PCR in symptomatic patients who are admitted as clinically suspect for COVID-19, while all other hospital patients who do not have symptoms suggestive of COVID-19 are tested by POCT. Patients in the latter category who test positive by POCT are re-tested by confirmatory PCR to exclude a false positive result.

This two-pronged algorithm (Figure 2) was based on the assumed performance characteristics and expected turnaround times of PCR and POCT. Since PCR was assumed to have greater accuracy but longer TAT, we reserved PCR testing for the higher prevalence group of symptomatic COVID-19 suspect patients. Using the faster but less sensitive POCT in this group would have a greater risk of false negatives, exposing health care providers, patients, and the community at large to undetected SARS-CoV-2. In asymptomatic patients, on the other hand, the pre-test probability of having the disease is low, so even with the less sensitive POCT there would only be a small number of false negatives. In fact, false positive results are of greater concern in this low-risk population, potentially leading to patient harm and misused resources in the form of unnecessary treatment, isolation, and PPE. Hence, we require PCR confirmation of all positive POCT results. Furthermore, we do not allow PCR-confirmed COVID-19 patients to be removed from isolation based on a negative follow-up POCT result. Based on local estimates, Table 2 exemplifies our rationale for using PCR in higher-prevalence symptomatic patients and POCT in lower-prevalence asymptomatic patients to minimize the risk of false negative results in both populations while at the same time optimizing turnaround time.

**Figure 2.**
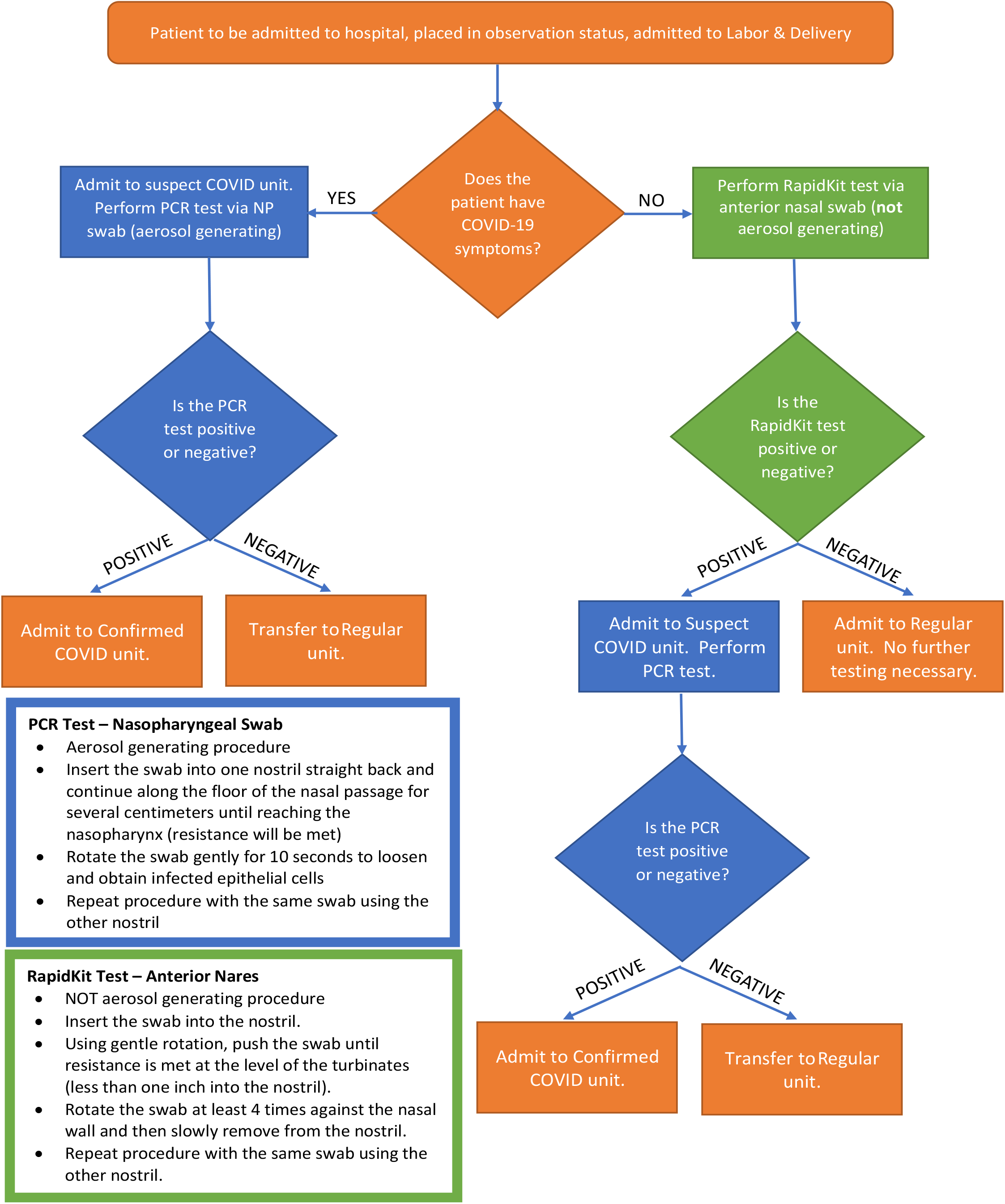
Our health care system guidelines for ordering PCR or POCT (locally known as RapidKit) in hospital patients.

**Table 2.**
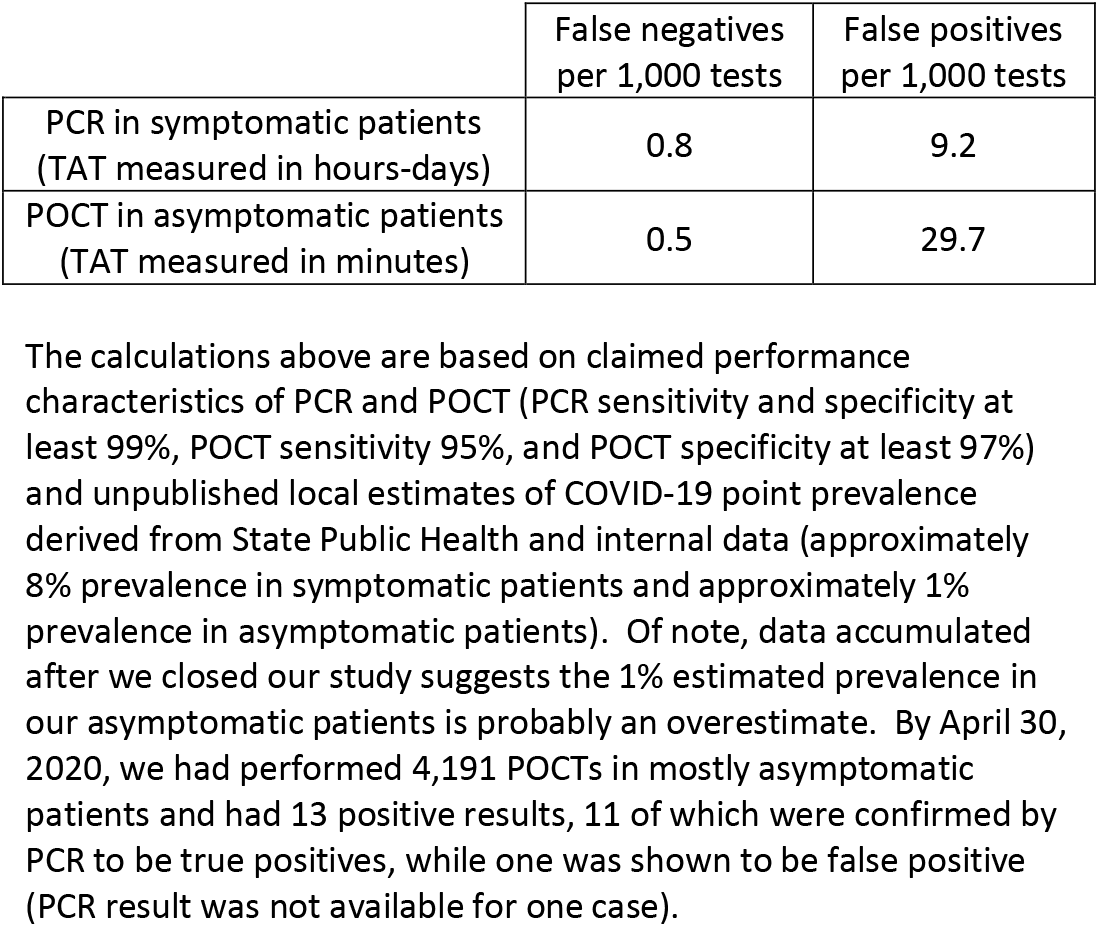
PCR and POCT false negative and false positive rates when utilized in different patient populations. The calculations above are based on claimed performance characteristics of PCR and POCT (PCR sensitivity and specificity at least 99%, POCT sensitivity 95%, and POCT specificity at least 97%) and unpublished local estimates of COVID-19 point prevalence derived from State Public Health and internal data (approximately 8% prevalence in symptomatic patients and approximately 1% prevalence in asymptomatic patients). Of note, data accumulated after we closed our study suggests the 1% estimated prevalence in our asymptomatic patients is probably an overestimate. By April 30, 2020, we had performed 4,191 POCTs in mostly asymptomatic patients and had 13 positive results, 11 of which were confirmed by PCR to be true positives, while one was shown to be false positive (PCR result was not available for one case).

Our algorithm for POCT was launched system-wide in April 2020, and after 3 weeks of implementation we had performed a total of 4,191 ID NOW COVID-19 tests in mostly asymptomatic patients. The experience has been successful in several aspects. At least 12 of 13 patients with positive POCT results appropriately received confirmatory PCR per the algorithm; 11 were confirmed by PCR while one was false positive. More importantly, POCT screening of asymptomatic hospital admissions during this time period identified 5 patients with SARS-CoV-2 infection who were not initially suspected of having COVID-19. Identification of these latent infections by POCT was undoubtedly beneficial to the care of our patients as well as safety of our hospital staff. Finally, to the best of our knowledge at the time of this writing there has not been a negative POCT-screened asymptomatic patient who was subsequently discovered to have COVID-19; while this is reassuring, its accuracy is uncertain without more rigorous investigation.

In summary, our experience with Abbott’s ID NOW COVID-19 assay does not refute the claimed sensitivity and specificity of the manufacturer. Although in our convenience sample of real world patients tested with both PCR and POCT we were not able to control for various factors (e.g., specimen type and quality, collection time relative to onset of symptoms, transport medium, time from collection to testing, etc.) the data we have accumulated to this point supports our two-pronged testing approach in which symptomatic hospital patients are tested by the more sensitive PCR method, while asymptomatic hospital patients with a low pre-test probability of infection are tested with POCT supplemented by PCR confirmation of positive results. Furthermore, in known COVID-19 patients who have been previously diagnosed by PCR we do not base deisolation decisions on a negative follow-up POCT result. We believe the results of this study may be applicable to similar health care systems that wish to standardize utilization of both PCR and POCT depending on local test availability.

## Data Availability

De-identified data is available upon request.

